# Viral shedding dynamics reveals sputum as a reliable and cost-saving specimen for SARS-CoV-2 diagnosis within the first 10 days since symptom onset: A prospective cohort study

**DOI:** 10.1101/2020.08.30.20183889

**Authors:** Jorge Levican, Leonardo I. Almonacid, Gonzalo Valenzuela, Tamara García-Salum, Luis Rojas, Eileen Serrano, Catalina Pardo-Roa, Erick Salinas, María José Avendaño, Fabiola Perazzo, Luis Antonio Díaz, Sebastián Valderrama, Marcos Ortega, Adriana Toro, Viviana Montecinos, Arnoldo Riquelme, Rafael A. Medina

**Affiliations:** Department of Pediatric Infectious Diseases and Immunology, School of Medicine, Pontificia Universidad Católica de Chile, Santiago, Chile; Department of Internal Medicine, School of Medicine, Pontificia Universidad Católica de Chile, Santiago, Chile; Program of Pharmacology and Toxicology, School of Medicine, Pontificia Universidad Católica de Chile; Department of Gastroenterology, School of Medicine, Pontificia Universidad Católica de Chile, Santiago, Chile; Department of Intensive Care Medicine, School of Medicine, Pontificia Universidad Católica de Chile, Santiago, Chile; Department of Respiratory Medicine, School of Medicine, Pontificia Universidad Católica de Chile, Santiago, Chile; Pediatric Service, Clínica UC San Carlos, Red Salud UC Christus, Faculty of Medicine, Pontificia Universidad Católica de Chile, Santiago, Chile; Department of Hematology-Oncology, School of Medicine, Pontificia Universidad Católica de Chile, Santiago, Chile; Department of Microbiology, Icahn School of Medicine at Mount Sinai, New York, USA

## Abstract

**Background:** Coronavirus disease 2019 (COVID-19) caused by the severe acute respiratory syndrome virus (SARS-CoV-2) is challenging global public health, due to an increasing demand for testing and the shortage of diagnostic supplies. Nasopharyngeal swab (NPS) is considered the optimal sample for SARS-CoV2 diagnosis and sputum (SPT) has been proposed as an economic alternative. However, the temporal concordance of diagnosis in NPS and SPT has not been addressed.

**Methods:** Through a longitudinal study we compared the shedding dynamics of SARS-CoV-2 RNA evaluated by RT-qPCR in serially collected SPT and NPS obtained from 82 ambulatory and hospitalized patients during acute infection and convalescence. The concordance during the follow-up and cost analysis between both collected specimens was evaluated.

**Findings:** We analyzed 379 samples, 177 NPS and 202 SPT. The highest proportion of positive samples was detected within the first 15 days after the symptoms onset. The median time of positivity was higher for NPS (median= 25 days) than SPT (median= 21 days). There was no significant difference in the median RT-qPCR C_T_ values between both sample types. The temporal categorization of matched-paired samples indicated substantial correlation (r=0·6023) and substantial agreement (87·23%) during the first ten days since symptoms onset (kappa = 0·697). A cost analysis demonstrated a significant saving when the SPT specimen was used.

**Interpretation:** Sputum is a feasible and cost-saving alternative to NPS, providing an equivalent value for the detection and follow-up of SARS-CoV-2 RNA.

**Funding:** Agencia Nacional de Investigación y Desarrollo (ANID) of Chile, NIH-NIAID USA.

## Introduction

In December 2019, a cluster of patients with pneumonia of unknown etiology and epidemiologically linked to a wet market in Wuhan, Hubei, China was described ^1^. The disease, known as COVID-19 has now widely spread in the world. To date a total of 24,997,327 cases and 842,522 deaths have been confirmed globally ^2^. The causative agent was found to be a *Betacoronavirus*, a virus closely related to the Severe Acute Respiratory Syndrome virus (SARS) that was named SARS-CoV-2 ^3^.

Nucleic acid screening tests were rapidly developed to detect the SARS-CoV-2 RNA and currently are widely used to diagnose COVID-19. The World Health Organization (WHO) recommends that the decision to test should be based on clinical and epidemiological factors, and the screening protocols should be adapted to the local situation ^4^. Although viral compartmentalization and shedding patterns of COVID-19 are not yet entirely understood, the NPS has been considered the optimal sample for the detection of SARS-CoV-2 nucleic acids ^4,5^. However, this procedure is invasive and produces discomfort in the patient during sample collection. Additionally, it frequently induces sneezing or coughing, being able to generate contaminated droplets with viral particles posing a significant risk to health workers ^6^. Of interest, recent studies have suggested that virus can also be detected for prolonged periods in saliva, sputum, and tracheal aspirates in patients with diverse clinical outcomes ^7-11^, suggesting that they might be an alternative sample for diagnosis of SARS-CoV-2 infection.

In addition, there is an increasing demand for flock swabs, commercial universal transport media, and personal protective equipment for health personnel worldwide ^12^. This shortage is becoming a worrisome perspective, particularly in low and middle-income countries. Hence, the availability of alternative sample collection protocols is urgent. Here, we demonstrate that SPT samples yield consistent results as compared to NPS samples for SARS-CoV-2 RT-qPCR detection, highlighting its use as an alternative sample for the diagnosis and follow-up of SARS-CoV-2 infection during the first ten days after the onset of the symptoms.

## Methods

### Patients and sample collection

A longitudinal study was conducted, including 82 patients diagnosed with COVID-19 in Santiago, Chile, between March 4th and July 30, 2020. The initial diagnosis was made by RT-qPCR of RNA extracted from a NPS performed at the Clinical Molecular Virology Laboratory of the UC-Christus Health network. Once COVID-19 diagnosis was confirmed, hospitalized or outpatients were invited to participate in the protocol. Written informed consent was obtained under protocol 16·066, which was reviewed and approved by the Scientific Ethics Committee at Pontificia Universidad Católica de Chile (PUC). Clinical and epidemiological data, along with clinical respiratory specimens (NPS and SPT) were serially collected by a trained nurse on days 1 to 36 after symptoms onset. NPS collection was conducted according to standard protocols. For SPT collection, patients were asked to voluntary cough and deposit the expectorated secretion in a sterile specimen container with 2 mL of saline phosphate solution. Samples were transported to the laboratory and processed the same day. The demographic and clinical characteristics of patients are described in Table 1.

**Table 1.** Demographic and baseline characteristics of patients infected with SARS-CoV-2.

|  | Outpatients<br>(n = 35) | Inpatients<br>(n = 45) | P value |
| --- | --- | --- | --- |
| **Characteristics** |  |  |  |
| Age, median years (range) | 33 (14-89) | **55 (0.75-83)** | 0.001* |
| **Sex** |  |  |  |
| Female, n (%) | 19 (54.3%) | 15 (33.3%) | 0.060 |
| Male, n (%) | 16 (45.7%) | 30 (66.7%) |  |
| **Symptoms** |  |  |  |
| **Respiratory** |  |  |  |
| Dyspnea, n (%) | 4 (11.4%) | **24 (53.3%)** | <0.001* |
| Chest discomfort, n (%) | 1 (2.9%) | 6 (13.3%) | 0.100 |
| Cough, n (%) | 24 (68.6%) | 37 (82.2%) | 0.155 |
| Runny nose, n (%) | 10 (28.6%) | 8 (17.8%) | 0.251 |
| Sore throat, n (%) | **13 (37.1%)** | 4 (8.9%) | 0.002* |
| **Constitutional** |  |  |  |
| Fever, n (%) | 27 (77.1%) | 31 (68.9%) | 0.412 |
| Chills, n (%) | **15 (42.9%)** | 6 (13.3%) | 0.003* |
| Headache, n (%) | **27 (77.1%)** | 11 (24.4%) | <0.001* |
| Myalgia, n (%) | **23 (65.7%)** | 16 (35.6%) | 0.007* |
| Severe fatigue, n (%) | 6 (17.1%) | **23 (51.1%)** | 0.002* |
| **Gastrointestinal** |  |  |  |
| Diarrhea, n (%) | 11 (31.4%) | 13 (28.9%) | 0.806 |
| Nausea/Vomiting, n (%) | 6 (17.1%) | 5 (11.1%) | 0.437 |
| **Sensorial** |  |  |  |
| Anosmia, n (%) | **21 (60%)** | 7 (15.6%) | <0.001* |
| Ageusia, n (%) | **14 (40%)** | 5 (11.1%) | 0.003* |
| **Comorbidities or conditions** |  |  |  |
| Obesity (BMI ≥ 30), n (%) | 6 (17.1%) | **17 (37.8%)** | 0.043* |
| Hypertension, n (%) | 4 (11.4%) | **14 (31.1%)** | 0.036* |
| Hyperlipidemia, n (%) | 3 (8.6%) | 10 (22.2%) | 0.101 |
| Prediabetes or NASH, n (%) | 2 (5.7%) | **16 (35.6%)** | 0.002* |
| Cardiovascular diseases, n (%) | 1 (2.9%) | 3 (6.7%) | 0.438 |
| Chronic pulmonary diseases, n (%) | 4 (11.4%) | 4 (8.9%) | 0.707 |
| Asthma, n (%) | 5 (14.3%) | 2 (4.4%) | 0.122 |
| Neurological disease, n (%) | 1 (2.9%) | 5 (11.1%) | 0.164 |
| Immuno-rheumatologic diseases, n (%) | 1 (2.9%) | 7 (15.6%) | 0.060 |
| Smoker, n (%) | 1 (2.9%) | 7 (15.6%) | 0.060 |
| Allergic disease, n (%) | **9 (25.7%)** | 3 (6.7%) | 0.018* |
| Duration of symptoms before admission, days (range) | - | 7.1 (1-15) | - |
| **Outcomes** |  |  |  |
| ICU admission, n (%) | - | 16 (35.6%) | - |
| 30-Day mortality, n (%) | 0 | 1 (2.2%) | 0.375 |

Abbreviations: BMI: Body mass index; NASH: Non-alcoholic steatohepatitis; ICU, Intensive care unit. \*p value <0.05, t-test/Chi-square test. Bold font highlight significant values associated to outpatients or inpatients. For two patients no clinical data were available.

#### RNA extraction and SARS-CoV-2 RNA amplification

The samples were subjected to RNA extraction using TRIzol LS^TM^ (Thermo Fisher Scientific), and SARS-CoV-2 RNA was detected using real-time one-step RT-qPCR targeting Orf1b with dual-labeled hydrolysis probe (6FAM/TAMRA) as previously reported with some modifications ^13^. Detailed protocol in Appendix 1.

#### RT-qPCR analytical validation

Identity of sequence of primers and probe regarding the virus circulating in Chile, linear range (Supplementary Figure 1), limit of detection, specificity and matrix interference (Supplementary Figure 2) were determined in the analytical validation of the test. Detailed protocols are described in Appendix 2.

### Statistical and cost analyses

Categorical parameters were expressed as frequencies (%), whereas continuous variables are presented as median (interquartile range [IQR]). Categorical characteristics comparison was performed with Chi-squared test. Agreement of the results between NPS and SPT specimens was assessed using Kappa statistics. According to Landis and Koch, Cohen’s kappa coefficient was defined as 0·00 - 0·20 = slight, 0·21-0·40 = fair, 0·41-0·60 = moderate, 0·61-0·80 substantial and 0·81−1 = almost perfect agreement ^14^ Kruskal-Wallis and Mann-Whitney U-test were used to evaluate differences between continuous variables, and Spearman’s correlation to assess the relation between NPS and SPT C**_T_**s. Kaplan-Meier estimator and Log-rank were used to evaluate differences in the positivity distribution over time. Wilcoxon matched-pairs signed rank test was used to compare matched C**T** pairs.

Cost analyses of collecting SPT versus NPS specimens were performed considering 1,000 samples. We evaluated one healthcare network provider (UC-Christus Hospital) and one private healthcare provider (private diagnostic laboratory). The cost of reagents was obtained directly from vendors or the inventory of the UC-Christus Hospital. We assessed the cost differences of obtention the SPT specimen as a self-collection procedure, with and without a health care worker assistance as compared to obtain NPS specimen, which requires a trained health care worker, a flock swab and a collection tube containing universal transport medium (UTM)(Table S1). A p-value <0·05 was considered statistically significant. Statistical analyses were conducted using PRISM 8·0 (v8-4·2).

### Role of the funding source

The funders had no role in study design; in the collection, analysis, and interpretation of data; in the writing of the report; and in the decision to submit the paper for publication. The corresponding author RAM, confirms that he had full access to all the data in the study and had final responsibility for the decision to submit for publication.

## Results

### SARS-CoV-2 RNA is detectable in SPT and NPS samples at comparable proportions during the early stages of COVID-19

Eighty-two patients were recruited for our longitudinal study (Table 1). Thirty-seven patients showed mild symptoms and did not require hospitalization, while 45 showed moderate to severe symptoms and were admitted to the hospital one to 15 days after the onset of symptoms. Of these, 35.6% required ICU and one patient died. For the inpatients, 66.7% were men and had a median age significantly higher than outpatients (55 vs. 33 years of age, respectively). Their principal clinical manifestations were dyspnea and severe fatigue. In outpatients the main clinical symptoms included sore throat, chills, headache, myalgia and the sensorial symptoms, anosmia and ageusia. Pre-existing conditions, including hypertension and prediabetes or non-alcoholic steatohepatitis (NASH) were more prevalent for inpatients and, allergic disease was more predominant for outpatients.

A total of 379 samples, 177 NPS, and 202 SPT were obtained (one to five samples were obtained per patient). Overall, 112 (63·28%o) NPS and 91 (45·05%) SPT samples were positive for SARS-CoV-2 and were distributed along the entire period of study, with the last positive sample detected in NPS at day 34 and at day 29 in SPT. In a detailed case-by-case analysis, we observed discontinuous viral RNA detection, i.e., after a negativization, the SARS-CoV-2 RNA was detected in a posterior sample. This dynamic was observed in ten of the 82 analyzed patients (12·2%), with six of them found only in NPS, three only in SPT and one in both sample types. Temporal stratification of SARS-CoV-2 RNA detection, showed that 77·27% of the samples were positive within the first five days post symptoms onset while the accumulated positivity reached 70·45% before 15 days (Table S1). To study the overall dynamics of viral RNA shedding in NPS and SPT over time, we used the Kaplan–Meier method. We observed a sustained decline in positivity during the time of the study, with no significant difference between the two sample types throughout the entire evaluated period (Log-rank test *p=*0·0623) (Figure 1). The same analysis showed that the median time of positivity (days at which the positivity in each group was 50%) was 21 days for SPT and 25 days for NPS. Similarly, when we separated the patients by severity, we observed a median time of positivity of 20 days for SPT and 23 days in NPS for outpatients (Log-rank test *p=*0·4671) and 21 days in SPT and 26 days in NPS for hospitalized patients (Log-rank test *p=*0·0650) (Supplementary Figure 3A and 3B). These data suggest that SARS-CoV-2 RNA excretion in NPS is more prolonged than SPT especially in a severe clinical context. Remarkably, we observed that RNA positivity in SPT was in high agreement with NPS during the first ten days. After this time, the positivity rate of both sample types shows a divergent pattern, suggesting a decrease in concordance (Figure 1A). A similar finding was obtained when the analysis was conducted separately in outpatients or hospitalized patients (Supplementary Figure 3A and 3B).

Subsequently, we compared the overall RT-qPCR C_T_ values of the positive NPS and SPT samples. The median C_T_ for NPS was 31.71 (IQR, 27·45-35·14) with a range of 13·6 to 38·87, and the median C**_T_** for SPT was 31·19 (IQR, 27·64-35·27) with a range of 16·5 to 38·07. No significant differences were found between the two groups (*p=*0·9108) (Figure 1B). The same was true when we analyzed the C**_T_** grouped by severity. The median C**_T_** for NPS was 32·63 (IQR 28·69-35·2) and for SPT was 33·17 (IQR 28·48-35·53) in hospitalized patients (*p=*0·6106), while for ambulatory patients the median was 30·78 (IQR 24·53-34·61) for NPS and 29·85 (IQR 23·9934·35) for SPT (*p=*0·7995). Although approximately two to three C**_T_**s higher were observed in the samples obtained from hospitalized patients than outpatients, this difference was not statistically significant (*p=*0·1431). As we expected, the highest viral loads (e.g. lower C**_T_** values) were found at the first days of the patients follow up (up to 10 days after symptoms onset). When we analyzed the C**T**s values over time, we observed a sustained decrease of viral load after the initial peak. Logistic regression analysis showed slopes of 0·5344 (95% CI= 0·3678-0·701) and 0·5685 (95% CI=0·4183-0·7187) for NPS and SPT respectively, with no significant differences (*p=*0-7640) (Figure 1C). While the Y-intercept was significant lower (*p=*0·0004) for NPS (29·14, 95% CI=26·17-32·11) than SPT (32·09, 95% CI= 29·36-34·82)(Figure 1C), this difference could be explained because of the prolonged shedding of viral RNA in NPS than SPT. This was reflected in the overall data set, where there are more abundant negative results (C_T_= or > to 39·02) in SPT than NPS, which resulted in an increased median C_T_ value of SPT. Thus, we conducted logistic linear regression including only the positive C_T_s (Figure 1D) for both sample types and we observed no significant difference in the slopes (0·3096, 95% CI=0·1804-0·4388 for NPS and 0·2890 95% CI=0·1337-0.4444 for SPT, *p=*0·8411) and the Y-intercept (26·17, 95% CI=24·15-28·18 for NPS and 27·09, 95% CI=24·92-29·26 for SPT *p=*0·3768). Overall, this suggests that the shedding dynamics in these two samples types are highly similar.

**Figure 1.**
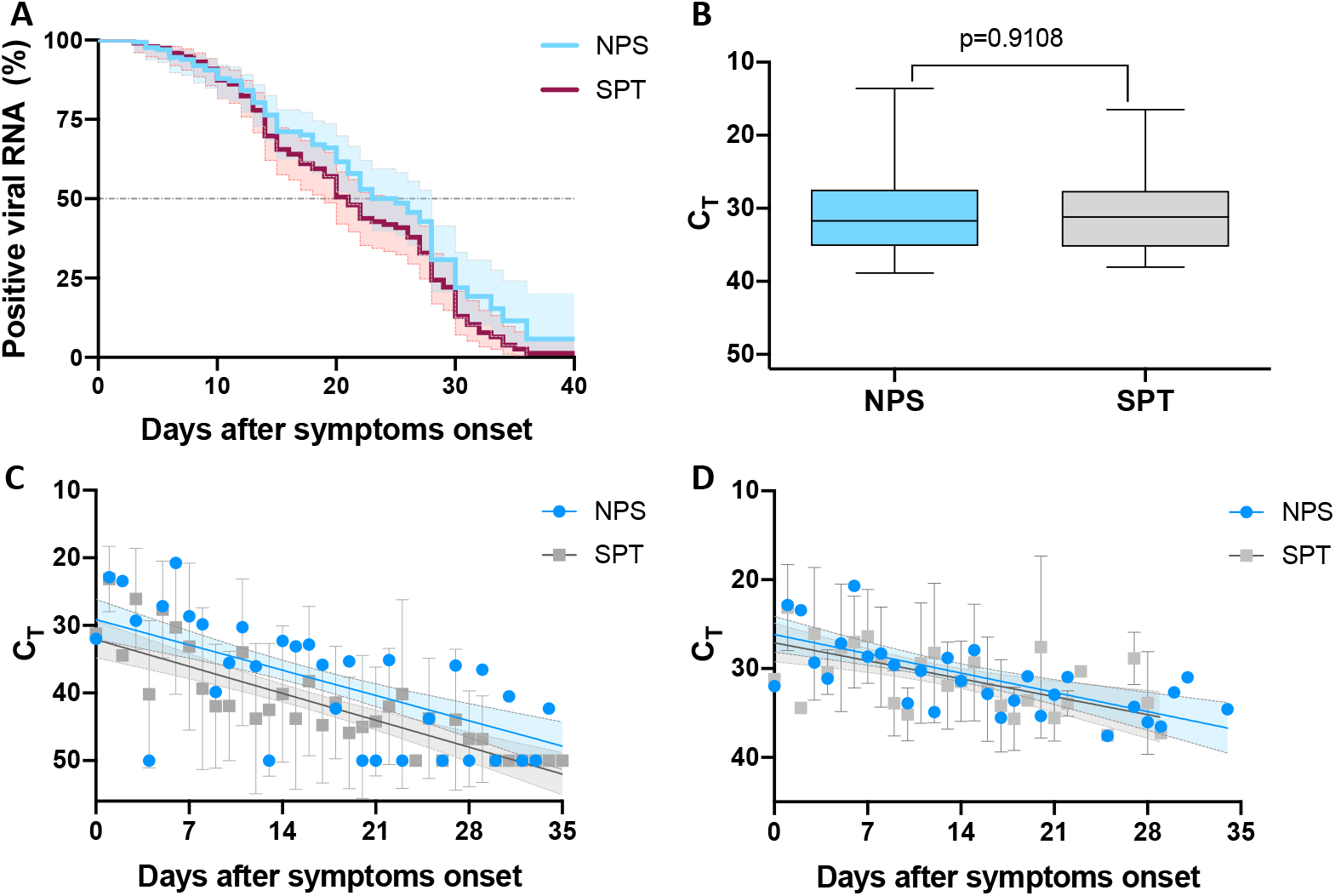
SARS-CoV-2 RNA is detectable in SPT and NPS samples at similar levels during the early stages of COVID-19. **A**. Percentage of SARS-CoV-2 RNA positive samples over time. Frequency of viral RNA detection was estimated using the Kaplan-Meier method. Log-rank (Mantel-Cox) test was applied to evaluate the difference in the duration of viral RNA shedding in the NPS (blue line) and SPT (red line) groups (95% CI are represented with the same colors) **B**. Overall comparison C_T_ from SARS-CoV-2 RNA positive samples. Median and range of C_T_s from NPS (n=112, blue box) and SPT (n=91, grey box) SARS-CoV-2 RNA positive cases. Non-parametric unpaired Mann-Whitney test was conducted to compare C_T_s from NPS and SPT. **C**. Temporal distribution of C_T_s of SARS-CoV-2 RNA of 177 NPS and 202 SPT (overall dataset) and D. 112 NPS and 91 SPT positive samples. Each data point represents the mean and error of the C_T_s obtained daily in a period of 35 days. NPS and SPT linear regression curve are shown in blue and grey respectively, and the 95% CI for each slope is shown in light blue (NPS) and light grey (SPT). The slopes and Y intercept were compared using *p* value using a two-tailed test.

### SPT has high concordance with NPS SARS-CoV-2 RNA detection during the first ten days of symptoms

To compare the agreement and correlation of NPS vs. SPT, in detecting SARS-CoV-2 RNA over time, 167 paired samples representing 34 days of follow up of 71 patients (30 outpatients and 41 hospitalized) were analyzed. One-hundred and three NPS (61·68%) and 76 SPT (45·51%) resulted positive for SARS-CoV-2 RNA, this data indicated a fair agreement (65·87%, Kappa=0·374, 95% CI 0·197-0·466) and overall Spearman’s correlation of r=0·4790 (95% CI=0·3489 to 0·5911, *p*<0·0001) (Table 2, Figure 2A).

**Table 2.** Analyses of paired NPS and SPT samples over time after symptoms onset.

| **Range (time)** | **Observed**<br>**agreement**<br>**NPS vs SPT** | **Kappa**<br>**value** | **SE**<br>**Kappa** | **95%CI** | **Agreement\*** |
| --- | --- | --- | --- | --- | --- |
| **1-7 days** | 89·29% | 0·661 | 0·144 | 0·309-1 | Substantial |
| **1-10 days** | 87·23% | 0·697 | 0·112 | 0·477-0·918 | Substantial |
| **11-34 days** | 57·60% | 0·177 | 0·083 | 0·016-0·339 | Slight |
| **1-34 days** | 65·87% | 0·331 | 0·066 | 0·197-0·466 | Fair |

\*According Landis and Koch <sup>14</sup>.

**Figure 2.**
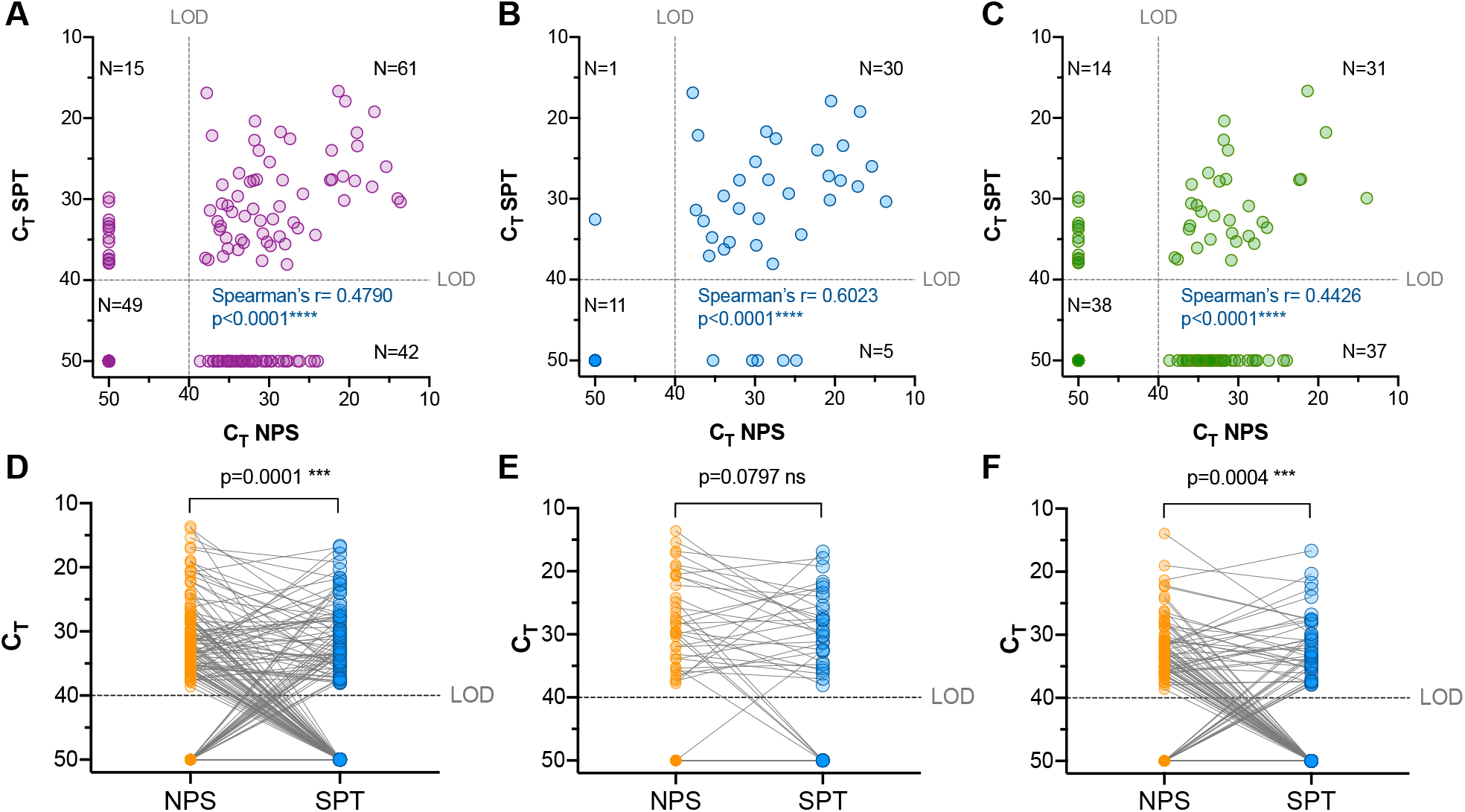
SPT has a higher correlation with NPS during the first 10 days of symptoms. **A**. Overall paired NPS vs SPT samples acquired during 36 days of follow-up (n = 167) were obtained from 30 mild and 41 hospitalized patients were plotted against each other. **B**. Paired NPS vs SPT paired samples (n = 47) obtained from 25 outpatients and 14 hospitalized patients during first ten days follow up after the symptom’s onset. **C**. Paired samples (n= 120) obtained from 25 outpatients and 37 hospitalized patients after ten days of the symptoms onset and up to 30 days of follow-up. Spearman’s correlation analyses results are shown on each graph. LOD= Limit of detection (C_T_<39·02 equivalent to 3·5 copies/µL). For graphical representation a C_T_ of 50 was assigned to negative samples. **D**. Overall matched pairs comparison of samples collected during the first ten days **(E)** and from days 11 to 36 **(F)** since onset of symptoms. Wilcoxon matched-pairs signed ranks test was used to compare the C_T_s obtained from NPS (orange dots) and SPT (blue dots) for the indicated time period.

Given that our results indicate an increased shedding of viral RNA during the first days after the symptoms onset, we stratified the data according to the temporal variable and conducted correlation and agreement analyses. The analysis showed a robust correlation in the early period of the study. In terms of positivity, the analysis of the first ten days after symptoms onset shows only one case of discrepancy for NPS and five for SPT with substantial to almost perfect agreement (87·23%, Kappa=0·697, 95% CI=0·477-0·918). In addition, when we compared the C_T_ values obtained with both sample types in this period we observed a strong correlation (*r*= 0·6023, 95% CI= 0·3736 to 0·7620, *p<* 0·0001) (Table 2, Figure 2B). Similar results were observed for the first ten days, and only moderate correlation and slight agreement was observed after ten days of symptoms onset (r=0·4426, Kappa=0·177 95% CI=0·016-0·339) (Figure 2C, Table 2). The same was true when we compared the C_T_ values from NPS and SPT in matched-paired samples collected simultaneously from the same individual. We found significant differences between the C_T_s of two sample types when the overall data set was compared (*p=*0·0001) (Figure 2D). Remarkably, when we compared paired C_T_s from the two sample types from 47 patients collected before ten days after symptoms onset, we found no significant differences between the groups (*p=*0·0797) (Figure 2E), while a significant difference was observed after ten days (*p=*0·0004)(Figure 2F). These data confirm that SPT is a feasible sample to detect SARS-CoV-2 and offers remarkable agreement with the results obtained with NPS sampling during the first ten days after the onset of COVID-19 symptoms.

### SPT specimen is a cost-saving alternative for the diagnosis of SARS-CoV-2 in a clinical setting

Assuming equivalent clinical efficacy between NPS and SPT within the first ten days since onset of symptoms, we developed a cost-minimization analysis to evaluate the feasibility of implementing SPT collection technique in the clinical practice. We estimated the direct and indirect medical costs associated with the labor hours of healthcare professionals and consumables associated with taking 1,000 specimens of each type of sample. The costs were based on 2020 economic variables and expressed in United States Dollars (USD). The economic data were obtained from a healthcare network setting and a private health diagnostic provider (Figure 3A). The procedure to obtain NPS specimen, which requires a trained health care worker, a flock swab and a UTM collection tube, was compared to the collection of a SPT sample collected by a health care worker or self-collected into a sterile container with saline solution as stabilizing medium. We found an overall cost reduction ranging from USD 2,340 to USD 5,478 per 1,000 specimens collected in both a health network setting and a private health diagnostic provider (Figure 3B). These analyses indicate that the collection of a SPT specimen provides a low-cost alternative for SARS-CoV-2 diagnosis.

**Figure 3.**
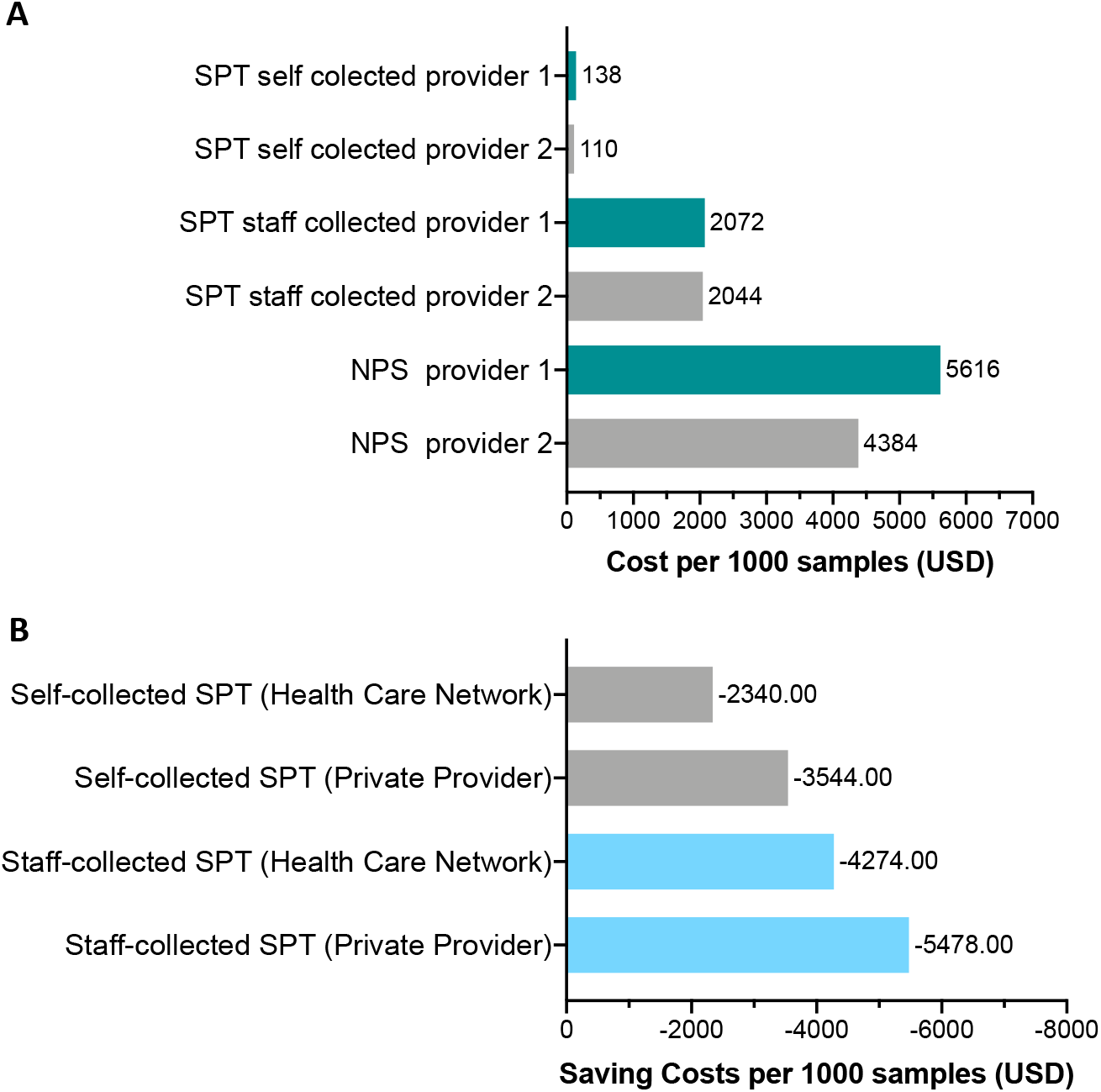
Comparison of costs of using SPT under clinical settings for diagnostics of SARS-CoV-2. **A**. Analysis of actual cost of obtaining SPT versus NPS samples through two different health network providers. **B**. Calculated cost reduction of collecting SPT versus NP specimens through a health network versus a private provider. Analyses performed considering 1,000 samples under different health services.

## Discussion

Currently there are scarce studies on the performance of different types of samples in the molecular diagnosing of SARS-CoV-2. Through a prospective cohort study, we demonstrate that SPT can be a valuable and cost-saving alternative sampling technique for COVID-19 diagnosis. By including individuals with a wide range of clinical manifestations, we show that this specimen can be used with an equivalent diagnostic yield as the standard NPS specimen, particularly within the first ten days since onset of symptoms, regardless of severity. Importantly, as the pandemic is now impacting the poorest regions of the world, our analyses suggest that this sampling technique can be easily applied at a fraction of the cost of the conventional NPS, providing an invaluable tool for community testing and surveillance in resource-limited countries.

Nasopharyngeal swab has been extensively used as the gold standard sample for molecular diagnosis of upper respiratory tract viral infections ^19^. In addition, other samples of the upper respiratory tree have been used for diagnosis, such as oro- and retropharyngeal swabs, saliva and sputum, and invasive samples such as bronchoalveolar lavage or tracheal aspirate have also been used ^20^. The choice of clinical specimen depends on the particular clinical condition of the individual and the clinical sensitivity of the test and the area sampled ^20, 19^. In the theory, the successful use of these samples ultimately is affected, in one hand, by the compartmentalization and timing of viral replication of each virus in the respiratory tract, and in the other hand, the timing of collection of the specimen. In the practice, the issue is obviously more complex and there is some level of discrepancy. Kim et al., described that adenoviruses and the endemic human coronaviruses are most frequently isolated from saliva, while influenza A and human rhinoviruses are isolated from NPS, concluding that no sampling method was consistently more sensitive than another ^21^. However, a different study found that sputum samples were significantly better than the NPS for the detection of influenza A, respiratory syncytial virus, human rhinovirus A, coronavirus OC43, and adenovirus in individuals with an acute respiratory infection ^22^.

Early on during the pandemic outbreak it was shown that SARS-CoV-2 RNA could be detected in bronchoalveolar lavage fluid (BLF), pharyngeal swabs, posterior oropharyngeal saliva, fibrobronchoscopy brush biopsies, stool, sputum, blood and nasal swabs in COVID-19 patients. Wang et al., observed that BLF (98%), sputum (72%) and nasal swabs (63%) were the samples that gave the most consistent results for viral RNA detection ^9^. In addition, a recent longitudinal study suggests that saliva samples are more sensitive for the detection of SARS-CoV-2 in COVID-19 hospitalized patients than NPS specimens ^17^ Similarly, To et al, (2020) in an observational COVID-19 cohort study of patients from two hospitals in Hong Kong, concluded that posterior oropharyngeal saliva samples are a non-invasive specimen more acceptable to patients and healthcare workers ^15^. In agreement, our study supports the use of SPT as an equivalent specimen to the NPS at early time points during infection.

These studies also showed that viral titers peak soon after symptom onset, and that higher viral loads were detected in the nose than the throat ^9,15,16^ In our study, we showed that viral RNA could be detected for up to 4 weeks (or more days in some patients) and there are frequent instances where individuals show discontinuous RNA detection, a phenomenon that was also observed in other studies ^11,15-17^ Although the precise timing and localization of viral replication requires more comprehensive research, recent evidence shows that angiotensin I converting enzyme 2 (ACE2 gene product), the receptor for the SARS-CoV-2 virus, is widely expressed in the respiratory tract and there is remarkably high expression level of ACE2 in secretory goblet cells and on nasal epithelial cells ^18^. This suggests that, at least in the first stage of the disease, high levels of replication could occur at these sites resulting in the detection of RNA from these samples ^18^.

Our cohort included individuals with a wide range of disease severity. This allowed us to determine that severity does not play a role in the detection of SARS-CoV-2 RNA in NPS and SPT samples by RT-qPCR, finding no significant differences in their diagnostic capacity during the first ten days after the onset of the symptoms. The longitudinal analysis of positive samples showed increased viral loads during the first week (lower C_T_s) and a gradual decrease over the following weeks, which was associated with the resolution of symptoms ^9,16^. Noteworthy, although no statistically significant, there was a slightly higher C_T_ (approximately two to three C_T_) values in hospitalized as compared to outpatients in both sample types. This difference could be explained because of more prolonged times of RNA excretion in hospitalized compared to ambulatory patients (the more delayed RNA excretion the higher the C_T_ at late time points). Accordingly, we observed higher median time of positivity in NPS for hospitalized patients (26 days) than ambulatory patients (23 days) while it was relatively constant in SPT (20 days for hospitalized vs 21 for ambulatory patients). Although not statistically significant, this data suggests that RNA excretion in NPS is more prolonged than SPT especially in a severe clinical context.

Our longitudinal study also allowed us to evaluate the correlation and consistency of using STP and NPS over time. We observed a gradual loss of correlation of NPS and SPT detection after ten days of follow up, which was concomitant with discontinuous detection of viral RNA in both sample types. We discarded analytical drawbacks that could be associated with matrix interference or loss of sensitivity in the RT-qPCR. In our study the NPS and SPT were collected by one trained nurse concurrently, transported immediately to the laboratory at 4°C, and processed on the day of collection. Hence, technical drawbacks related to the sample collection are unlikely to explain this loss of correlation seen after the ten days. We hypothesized that our ability to detect viral RNA with the different specimens could be a reflection of the timing of replication of the virus in the nasopharyngeal cavity versus the other respiratory tract sites during COVID-19 disease. It is plausible that after ten days in most cases the virus could be replicating at low levels and only in restricted areas of the respiratory tract. Alternatively, we could be detecting traces of SARS-CoV-2 RNA remaining and not necessarily active viral replication. This agrees with previous reports showing that the virus can be isolated in culture only within the first week after the symptoms onset in sputum ^11^. Overall, our findings indicate that the timing and sampling site must be carefully considered for diagnosis and there is a need to design cohort studies to further characterize the natural infection of COVID-19 patients longitudinally.

The COVID-19 pandemic has expanded rapidly worldwide and currently Latin America has become the epicenter of the pandemic, stressing health systems and causing large economic losses^25-27^. SARS-CoV-2 is highly transmissible in the general population, which has highlighted the need to improve active surveillance, early detection, isolation and tracking of COVID-19 cases as a fundamental part of control strategies ^30^. The ease of obtaining a SPT sample, which can be self-collected, together with our costs analysis demonstrated that the use of this sample type is a cost-saving and suitable alternative for SARS-CoV-2 detection. Importantly, we show that it has a similar diagnostic value as NPS when obtained within the first ten days since of symptom onset. In addition, the use of alternative sample collection, such a SPT specimens, should be included in the context of active surveillance and in the clinic upon confirming the timing of onset of symptoms. Overall, this strategy can help relieving some economic, logistic and biosecurity constraint associated to the use of NPS.

## Data Availability

All data presented in this manuscript are available to other researchers upon request to the corresponding researcher

## Contributors

J.L. and L.I.A. designed experiments, acquire and analyzed the data, prepared illustrations, and wrote the paper. G.V., F.P., T.G.S., M.J.A obtained and analyzed clinical metadata, prepared tables and wrote the paper. L.R. analyzed the data, prepared illustrations, and wrote the paper. Ei.S. and C.P. analyzed the data and wrote the paper. Er.S, L.A.D., S.V., M.O., A.T. recruited patients and collected clinical metadata and revised the paper. V.M. provided funding and resources and wrote parts of the paper. A.R. recruited patients and collected clinical metadata, analyzed the data and wrote the paper. R.A.M. obtained funding, conceived and supervised this study, designed experiments, analyzed the data, prepared illustrations, and wrote the paper.

## Declaration of interest

We declare no competing interest.

## Acknowledgment

Protocols and the study set-up used for this study were based on influenza virus studies established in part with the support of the FONDECYT 1161971 and ACT 1408 grants from the Agencia Nacional de Investigación y Desarrollo (ANID) from Chile, the FLUOMICS Consortium (NIAID grant U19AI135972) funded by NIH and with funding from CRIP (Center for Research on Influenza Pathogenesis), an NIH funded Center of Excellence for Influenza Research and Surveillance (CEIRS, contract number HHSN272201400008C) to RAM. Special thanks to Leandro Fernandez García, for technical assistance in the preparation of synthetic SARS-CoV-2 RNA standard.

## Notes

### Competing Interest Statement

The authors have declared no competing interest.

### Author Declarations

Written informed consent was obtained under protocol 16-066, which was reviewed and approved by the Scientific Ethics Committee at Pontificia Universidad Catolica de Chile (PUC)

